# Pulmonary alveolar regrowth in an adult COVID-19 patient

**DOI:** 10.1101/2020.05.10.20097634

**Authors:** Jingyu Chen, Huijuan Wu, Yuanyuan Yu, Nan Tang

## Abstract

We detected active alveolar regrowth in the lung of a 58-year-old COVID-19 patient who underwent lung transplantation due to severe lung hemorrhage. Specifically, immunohistological and scanning electronic microscopy analyses revealed that alveolar type II epithelial cells (AT2 cells) accumulate in response to viral pneumonia and that these AT2 cells actively proliferate and differentiate into squamous AT1-like alveolar epithelial cells. Thus, our work establishes that alveolar regrowth does occur in post-COVID-19 injury adult human lungs.

---

Alveolar regrowth after an acute lung injury has been observed in many mammals. However, alveolar regrowth after acute lung injury in adult humans is still poorly characterized, mainly due to the lack of lung samples and regeneration-specific molecular markers. Since December 2019, COVID-19, an infectious disease caused by the severe acute respiratory syndrome coronavirus 2 (SARS-CoV-2), is affecting more than 200 countries. Although many confirmed cases of asymptomatic infection or mild upper respiratory illness have been resolved, some cases continue to develop into severe viral pneumonia (Bai et al., 2020; Wang et al., 2020). Both our basic understanding of the COVID-19 disease and our ability to prognosticate patient outcomes would be substantially improved if we knew whether alveolar regrowth occurs after viral-pneumonia-related diffuse alveolar injuries.

The alveolar epithelium comprises two types of alveolar epithelial cells: alveolar type I (AT1) and alveolar type II (AT2) cells. Work in animal models has shown that AT2 cells function as resident alveolar stem cells that can proliferate and differentiate into AT1 cells to build new alveoli after lung injury (e.g., lung lobe resection or chemicals, virus, or bacteria insults) (Hogan et al., 2014; Katsura et al., 2019; Khatri et al., 2019; Liu et al., 2016; Rock et al., 2011; Wu et al., 2020). However, in patients with COVID-19 pneumonia, SARS-CoV-2 can directly attack alveolar epithelial cells and cause massive AT2 cell death. It is unknown whether AT2 cells contribute to post-SARS-CoV-2 infection alveolar regrowth.

The COVID-19 patient is a 58-year-old male, who is a non-smoker without chronic diseases except he is a hepatitis B carrier. Prior to the infection, the patient did not have signs of lung disorders. Starting with admission, this patient received oxygen support to treat his hypoxemia. In the course of the disease, noninvasive ventilation, intubation, invasive ventilation, and extracorporeal membrane oxygenation (ECMO) were used in succession (Fig. S1a) (Chen et al., 2020). However, hemorrhage in the lungs occurs on disease onset day 36 (Fig. S1a). Therefore, an emergency lung transplant was performed on disease onset day 38.

H&E staining of the lung specimen from the COVID-19 patient revealed pulmonary interstitial thickening, hemorrhage, and fibrotic changes (Fig. 1a). Multiple cell aggregates and hyaline membranes were still evident in the alveolar lumen, indicating severe diffuse alveolar damage (Fig. 1a). Immunostaining showed signs of lung fibrotic changes, including significant collagen I deposition and proliferating α-SMA myofibroblasts (Fig. S2a, b). To investigate the proliferation of AT2 cells, we performed immunostaining experiments using antibodies against HTI-56 (a marker of human AT1 cells), Prospc (a marker of AT2 cells), and Ki67. In most regions of the COVID-19 lung, very few HTI-56^+^ AT1 cells were observed, indicating a significant depletion of AT1 cells (Fig. 1b). Notably, we observed that alveolar regions harboured a large number of clustered AT2 cells lining the alveolar epithelium in the COVID-19 lung (Fig. 1b). About 1.1% of these AT2 cells stained positive for Ki67, indicating that AT2 cells are proliferating (Fig. 1b). These findings establish that AT2 cells are able to replicate additional AT2 cells after SARS-CoV-2 induced lung injury.

**Fig. 1.**
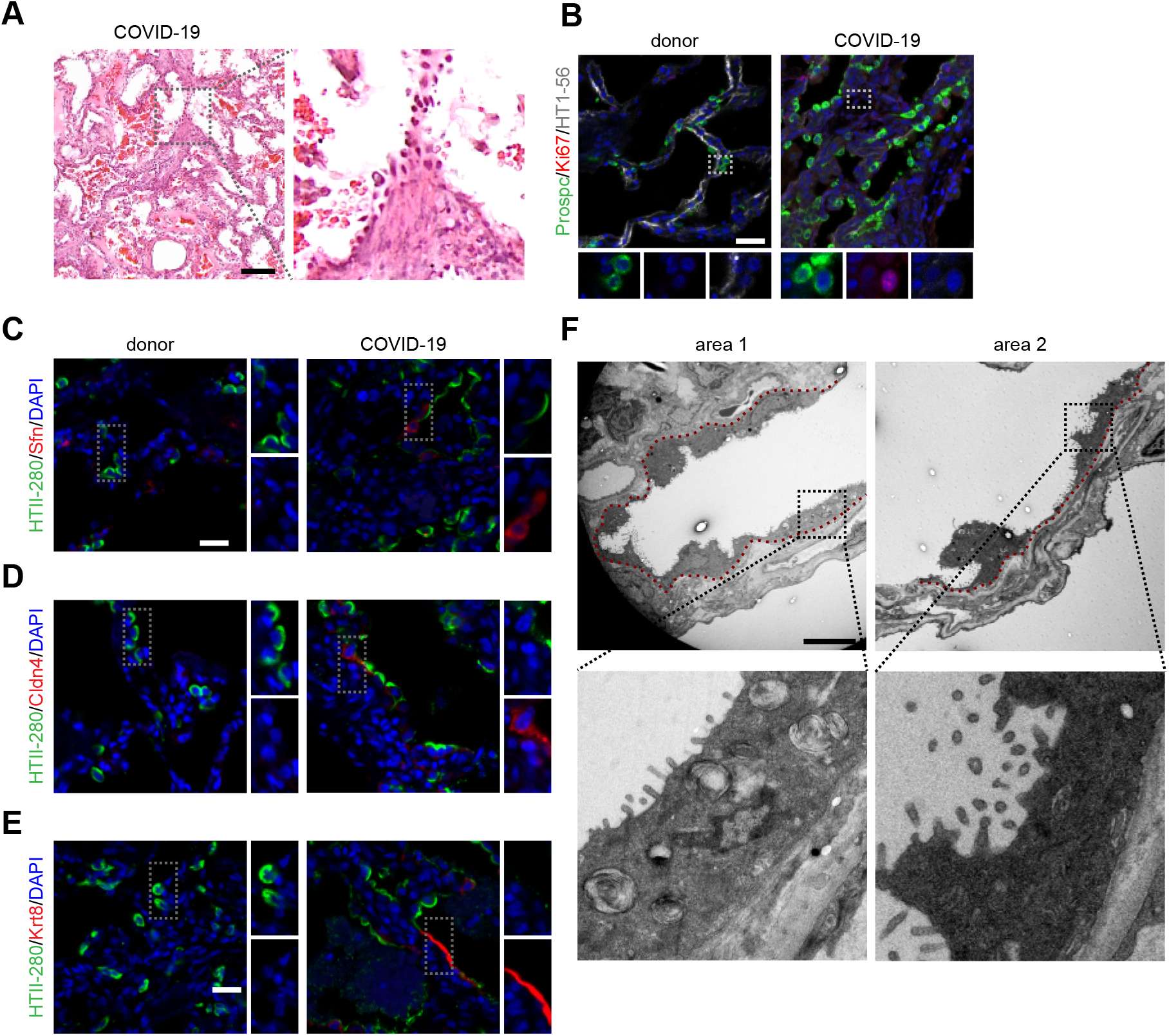
The proliferation and differentiation of the resident alveolar stem cells in the COVID-19 lung. **a**. H&E staining of the COVID-19 lung shows lung fibrosis and severe lung injury. **b**. Immunostaining using antibodies against Prospc, Ki67, and HTI-56 of a healthy donor lung and the COVID-19 lung. **c-e**. Immunostaining using antibodies against HTII-280 and Sfn (**c**), HTII-280 and Cldn4 (**d**), and HTII-280 and Krt8 (**e**) of a healthy donor lung and the COVID-19 lung. **f**. The alveolar epithelium of multiple regions of the COVID-19 lung is lined by AT1-shape like AT2 cells. The red dashed line indicates the location of basement membrane. Scale bars, 100μm (a), 20μm (b-e), 5μm (f).

Previous studies in both human and mouse lungs have confirmed the existence of a transient intermediate cell state for AT2 cells that occurs during differentiation of AT2 cells into AT1 cells (Jiang et al., 2020; Kobayashi et al., 2019; Riemondy et al., 2019; Strunz et al., 2019; Wu et al., 2020). AT2 cells in this intermediate cell state subsequently differentiate into AT1 cells (Jiang et al., 2020; Kobayashi et al., 2019; Riemondy et al., 2019; Strunz et al., 2019; Wu et al., 2020). We used immunostaining to examine the COVID-19 lung and a healthy donor lung with three markers with transient expression profiles known to specifically define this intermediate AT2 cell state: Claudin4 (Cldn4), Stratifin (Sfn), and Keratin 8 (Krt8). None of the AT2 cells in the healthy donor lung expressed any of these markers. In contrast, many HTII-280^+^ AT2 cells in the COVID-19 lung expressed Cldn4, Sfn, or Krt8 (Fig. 1c-e). Some AT2 cells that express Cldn4, Sfn, or Krt8 show decreased expression levels of HTII-280 and Prospc (Fig. S3a). Collectively, these observations confirm that AT2 differentiation was initiated in the COVID-19 patient’s lung by the 38th day of disease onset.

Given the knowledge that AT2 intermediated cell state cell markers are transiently expressed in differentiating AT2 cells but not expressed in AT1 cells, we next used scanning EM to investigate the potential differentiation of AT2 cells (Fig. 1f). Consistent with our immunostaining results that show a significant depletion of AT1 cells in the COVID-19 lung (Fig. 1b), few AT1 cells were observed by the scanning EM analysis. However, the scanning EM images showed that multiple regions typically occupied by AT1 cells in normal lungs actually harboured numerous squamous alveolar epithelial cells (Fig. 1f). Although these cells did contain some AT2-cell-like features—including lamella bodies and distinct apical microvilli—they exhibited the characteristic flattened and elongated shape of AT1 cells (Fig. 1f). The expression of the intermediated AT2 cell markers can be also detected in AT2 cells of IPF lungs (Jiang et al., 2020; Kobayashi et al., 2019; Strunz et al., 2019), however, no squamous and elongated AT2 cells can be observed in IPF lungs by scanning EM analysis, supporting the hypothesis that AT2 cells in IPF patients may be impaired in further differentiation into AT1 cells (Jiang et al., 2020; Wu et al., 2020). Together, these findings suggest that, in addition to the AT2 cell proliferation and differentiation into the intermediate AT2 cell state that we observed, the AT2 cells of the COVID-19 patient lung can apparently participate in alveolar regrowth by differentiating into AT1-like cells. Additionally, our results establish that alveolar regrowth can be activated by humans in their 50s.

An interesting remaining question concerns the time scale of the alveolar regeneration process in adult human lungs. In rodents, it takes a few weeks to regenerate the alveoli and restore lung functions (Barkauskas et al., 2013; Desai et al., 2014; Liu et al., 2016; Zacharias et al., 2018). To date, there is no experimental evidence that new alveolar structures can be created in the adult human lung. One medical study showed that a 33-year-old woman who had undergone pneumonectomy had alveolar growth over a period of 15 years; this study relied on MRI-based assessment of lung microstructure and analyses of lung function (Butler et al., 2012). For most patients who have recovered from COVID-19 pneumonia, CT scans indicate that the extent of lung disease gradually recover during the third week post symptom onset. Our results demonstrate that alveolar regrowth in COVID-19 lungs was initiated by (at the latest) the 38th day after the onset of initial symptoms. These observations indicate that it may take a few weeks for AT2 cells to repopulate and to differentiate into AT1 cells in adult humans after acute lung injuries. Future imaging studies that analyze the lung microstructure of patients who recovered from viral pneumonia should provide further insights into whether new functional alveolar structures will form after acute lung injury.

Pulmonary fibrosis is a frequent complication in patients with viral pneumonia-induced acute respiratory distress syndrome (ARDS). However, CT scans have shown that the signs of pulmonary fibrosis after viral pneumonia can partially regress over time (Toufen Jr et al., 2011). Although the patient in the present study underwent lung transplantation surgery due to severe lung hemorrhage, it seems plausible that alveolar regrowth may also occur in patients with less severe viral pneumonia, and to speculate that such regrowth functions to help to restore lung function and even resolve pulmonary fibrosis. Our observations open the door for future studies to elucidate the mechanisms that regulate human alveolar regeneration after acute lung injury and facilitate to prognosticate the outcomes of COVID-19 patients.

**Fig. S1. The medical history of the COVID-19 patient.**

**a**. A summary of the medical history of the COVID-19 patient. **b**. Illustration of explanted lungs from the COVID-19 patient.

**Fig. S2. The COVID-19 lung shows fibrotic changes.**

**a**. Immunostaining using antibodies against HTII-280 and Collagen I of a healthy donor lung and the COVID-19 lung. **b**. Immunostaining using antibodies against α-SMA and Ki67 of a healthy donor lung and the COVID-19 lung. Scale bars, 20μm (a, b).

**Fig. S3. Some AT2 cells at the intermediate cell state show reduced levels of AT2 markers.**

**a**. Some Krt8 expressing AT2 cells (arrowhead) show reduced expression levels of HTII-280 (green) or Prospc (grey).

## MATERIALS AND METHODS

| REAGENT | SOURCE | IDENTIFIER |
| --- | --- | --- |
| Antibodies |  |  |
| Rabbit anti-Collagen I | Abcam | Cat# ab34710 |
| Rabbit anti-Prospc | Millipore | Cat# AB3786 |
| Mouse anti-α-SMA | Sigma | Cat# C6198 |
| Rat anti-Ki67 | Invitrogen | Cat# 14-5698-82 |
| Mouse anti-HTI-56 | Terrace Biotech | Cat# TB-29AHT1-56 |
| Mouse anti-HTII-280 | Terrace Biotech | Cat# TB-27AHT2-280 |
| Rat anti-Krt8 | DSHB | Cat# TROMA -I |
| Rabbit anti-Cldn4 | Invitrogen | Cat# 36-4800 |
| Rabbit anti-Sfn | Invitrogen | Cat# PA5-95056 |
| Alexa Fluor 568 Donkey anti-rabbit | Thermo Fisher Scientific | Cat# A10042 |
| Alexa Fluor 647 Donkey anti-mouse | Jackson Immuno Research | Cat# 715-605-151 |
| Alexa Fluor 488 Donkey Anti-Rat | Jackson Immuno Research | Cat# 712-545-150 |
| Biotin-Donkey Anti-Rabbit | Jackson Immuno Research | Cat# 711-065-152 |
| Biotin-Donkey Anti-Rat | Jackson Immuno Research | Cat# 712-065-153 |
| Reagent |  |  |
| Hemotoxylin | Abcam | Cat# ab150678 |
| Eosin | Sigma | Cat# HT110280 |
| Tyramide | Perkin Elmer | Cat#NEL744001KT |

### Human specimens

All experiments with human tissue samples were performed under protocols approved by the Institutional Review Boards at National Institute of Biological Sciences, Beijing and Wuxi People’s Hospital. The COVID-19 lung tissues were collected from lung explants of the COVID-19 patient (male, age 58yr). The healthy donor lung tissues (male, age 56yr) were resected on the basis of size incompatibility.

### Hematoxylin and Eosin (H&E) staining

Lungs were fixed with 10% neutral buffered formalin, cryoprotected in 30% sucrose and embedded in OCT, finally cut into 15μm thick cryosections. The H&E staining experiment followed the standard H&E protocol.

### Immunostaining experiments

The human lung tissues were fixed with 10% neutral buffered formalin, cryoprotected in 30% sucrose, and embedded in OCT. 15μm thickness cryosections were used for immunostaining experiments. The primary antibodies used were as follows: rabbit anti-Collagen I (1:300), mouse anti-a-SMA (1:300), rabbit anti-Prospc (1:500), mouse anti-HTII-280 (1:50), mouse anti-HTI-56(1:50), rat anti-KI67 (1:200), rabbit anti-CLDN4 (1:500), rabbit anti-SFN (1:500), rat anti-KRT8 (1:200). All of the Alexa Fluor coupled secondary antibodies were used at 1:500 dilutions. Nuclei were stained with DAPI. The slides were mounted with 50% glycerol. For the KRT8, CLDN4 and SFN staining experiment, the tyramide (Perkin Elmer, NEL744001KT) signal amplification method was used followed the manufacturer’s recommendations.

### Transmission electron microscope

The human lung tissues were fixed with a mixture of 4% PFA and 0.4% glutaraldehyde for 24 hours at 4°C. Lungs were cut into small pieces (about 1mm^3^) and soaked in 1% antimony tetraoxide for 1 hour at 4°C. Then the tissues were dehydrated in acetone, and embedded by resin. The embedded resin blocks were cut into ultra-thin sections and further stained using uranium acetate and lead citrate. The TEM pictures were taken by a transmission electron microscopy (FEI, Tecnai spirit G2).

## Data Availability

All data are included in the manuscript.

## ACKNOWLEDGEMENT

This work was supported by grants from the National Key Research and Development Program of China (SQ2020YFA070010) and Foundation for Special Projects of COVID-19 Prevention and Control in Wuxi city (No2020X002).

